# An Integrated Multi-omic Analysis Reveals Novel Gene-Metabolite Relationships in Human Steatohepatitic Hepatocellular Carcinoma

**DOI:** 10.64898/2026.01.28.26344977

**Authors:** Garrett B. Anspach, Robert M. Flight, Sehyung Park, Hunter N. B. Moseley, Robert N Helsley

## Abstract

**Background:** Metabolic dysfunction-associated steatotic liver disease (MASLD) is the fastest-growing etiology of hepatocellular carcinoma (HCC). A mechanistic understanding of the metabolic heterogeneity of MASLD-driven tumors is crucial to inform strategies for future treatment options.

**Methods:** Paired tumor (n=8) and adjacent non-tumor tissue (n=8) were collected from patients with steatohepatitic HCC at the University of Kentucky Markey Cancer Center. Hematoxylin and eosin (H&E) staining was used for pathological determination of tumor and adjacent nontumor tissue by a board-certified pathologist. Lipidomic, metabolomic, and transcriptomic analyses were performed, and data were integrated across platforms to identify novel relationships across tumor and adjacent nontumor tissue.

**Results:** Histological analysis by H&E showed significant lipid vacuole accumulation and inflammatory foci in HCC tumors relative to nontumor tissue. Across omics platforms, we identified 1,679 genes, 1,696 metabolites, and 292 lipids that were significantly (padj<0.01) increased or decreased in tumors relative to nontumor tissue. We identified significant reductions in total ceramides and increases in fatty acyl chain saturation in tumor tissue. Furthermore, metabolites involved in amino acid and fatty acid metabolism were largely decreased in tumors relative to nontumor tissue. We also identified a total of 303 highly significant and novel transcript-metabolite associations (117 gene-metabolite; 186 gene-lipid) across tumor and nontumor tissue.

**Conclusions:** Taken together, this integrative analysis reveals novel relationships between steady-state gene transcripts and specific metabolites in steatohepatitic tumors, thereby identifying new pharmacological targets that may be exploited for therapeutic benefit.

## INTRODUCTION

Hepatocellular carcinoma (HCC) is the most common form of liver cancer worldwide, accounting for ∼90% of all liver-related cancers (1). Current projections estimate that greater than 1 million individuals are affected by HCC annually (1). The prognosis of HCC associates closely with tumor stage, with early stage diagnoses associating with greater 5-year survival rates at ∼70%, while more advanced HCC associating with rates as low as ≤20% (2, 3). Survival rates also depend on tumor heterogeneity, etiology, treatment, and patient comorbidities (4). Viral-induced hepatitis remains the foremost risk factor for HCC; however, the prevalence of metabolic dysfunction-associated steatotic liver disease (MASLD) is rapidly escalating and emerging as a prominent etiology of HCC. In the United States from 2004-2009, MASLD-driven HCC showed a 9% annual increase with ∼14% of all HCC cases being attributed to MASLD (5). Across Asian countries, it is estimated that HCC will increase by 44-85% from 2016 to 2030 in large part due to non-viral factors, such as MASLD and its comorbidities (obesity, diabetes, hyperlipidemia) (6). This estimated global shift in HCC etiology from viral to non-viral sources warrants identification of novel pathways in MASLD-HCC pathogenesis that may be exploited for therapeutic benefit.

The World Health Organization approximates that 35% of all HCC cases can be histologically classified into one of eight subtypes. The most common subtype is Steatohepatitic Hepatocellular Carcinoma (SH-HCC), accounting for up to 20% of all cases (7, 8). Histologically, SH-HCC often reflects that of MASLD and metabolic dysfunction-associated steatohepatitis (MASH) in the nonneoplastic liver, including the presence of steatosis, cell ballooning, inflammation, fibrosis, and Mallory-Denk bodies (7, 8). The SH-HCC variant was diagnosed in ∼36% of patients with confirmed MASH or MASLD with increased alcohol intake (MetALD), as compared to ∼1% of individuals with other liver disease etiologies (9). Consistent with a MASLD-driven etiology, approximately 50% of SH-HCC patients had at least 3 metabolic syndrome components, as compared to 22.5% with other etiologies (9). Despite the increased associations with metabolic syndrome, 5-year survival rates of patients with SH-HCC are comparable to that of patients with non-SH-HCC (10). Other unique histological features of SH-HCC, as compared to conventional HCC, include the number of activated stellate cells and subsequent fibrosis within the tumor core, as well as loss of cytokeratin 8/18 expression within ballooned hepatocytes of SH-HCC (9). While SH-HCC represents an increasingly prevalent subtype of HCC, a mechanistic understanding of the metabolic heterogeneity of SH-HCC tumors is crucial to inform strategies for future treatment options.

In this manuscript, we collected eight paired tumor and adjacent nontumor tissue from patients with biopsy-confirmed HCC who met at least one of five cardiometabolic risk factors or who met the cutoff for MetALD (11), and exhibited a tumor microenvironment consistent with that of SH-HCC by histological analysis. We then employed omics-based techniques to identify novel relationships between RNA transcripts, lipids, and metabolites in these paired tumor and adjacent nontumor tissue. We report significant alterations in lipid saturation, and amino acid and fatty acid transport and metabolism, with a total of 303 highly significant transcript-metabolite associations identified across paired tissue. This methodological approach offers in-depth pairwise analyses of integrated data across omics platforms, thereby aiding in the identification of novel pathways controlling cancer progression.

## METHODS

### Human tissue collection

Patients undergoing surgical resection of primary HCC at the University of Kentucky (UK) were consented for tissue donation under institutional review board protocols #44026 and #70872. Surgical resection specimens were collected as part of routine patient care and sent by the surgeon to UK surgical pathology for dissection and assessment. Tumor and non-tumor adjacent tissue were then provided to the Biospecimen Procurement and Translational Pathology (BPTP) Shared Resource Facility of Markey Cancer Center staff. Samples were aliquoted, flash frozen, and then stored in vapor-phase liquid nitrogen freezers in the BPTP. Aliquots were transferred on dry ice to the BPTP histology laboratory, embedded in OCT, and sectioned for hematoxylin and eosin (H&E) staining. The H&E slides were reviewed by an independent pathologist to identify tumor versus adjacent non-tumor tissue. Deidentified tissue samples were then provided to the researchers for bulk RNA-sequencing, lipidomic, and metabolomic analyses. An experimental overview is provided in **Supplemental Figure 1**.

### Bulk RNA-Sequencing

RNA was isolated from paired tumor and adjacent nontumor tissue via the RNeasy Mini kit (Qiagen). The quantity and quality of the samples were determined using the Cytation 5 (BioTek) plate reader and Agilent 4150 Tape Station System, respectively. RNA that did not meet an RNA integrity cutoff of 4.0 were not included in the analysis (**Supplemental Table 1**). mRNA-seq libraries and paired-end sequencing were all performed by Novogene, as previously described (12, 13).

### University of California (UC) West Coast Metabolomics

Tissue sample extractions, lipidomic and metabolomic analyses were performed by the UC Davis West Coast Metabolomics Center. Extraction for lipids and polar hydrophilic small molecules were performed using a previously established methyl-tert-butyl ether (MTBE) protocol (14). In short, 4 mg of liver tissue was weighed and 225 μL of ice-cold methanol containing an internal standard mixture was added with 750 μL of MTBE. Samples were then vortexed for 10s and mixed by shaking for 5 min at 4°C. After, 188 μL LC-MS grade H2O was added, samples were then vortexed for 20 s and centrifuged for 2 min at 14,000 x rpm for phase separation. The organic (upper; 350 μL) phase and the aqueous (bottom; 110 μL) phase were separated into individual tubes and dried down for reverse-phase liquid chromatography-high resolution tandem mass spectrometry (LC-HRMS/MS) for lipidomics, hydrophilic interaction chromatography (HILIC) LC-HRMS/MS for biogenic amines, and gas chromatography-time-of-flight mass spectrometry (GC-TOFMS) for the primary metabolism platform.

### Lipidomics

After dry down, samples were reconstituted in 110 μL MeOH:Tol (9:1) + 12-cyclohexylamino-carbonyl-amino-dodoecanoic acid (CUDA; 50 ng/mL) + Avanti SPLASHone lipidomics internal standard mixture. Samples were vortexed for 10s, sonicated for 5 min, and centrifuged for 2 min at 16,000 x *g*. Fifty μL was transferred to two separate glass amber vials with microinserts for positive and negative ionization modes.

#### Positive ionization mode

Mobile phase consists of A: 60:40 ACN:H_2_O+10mM ammonium formate+0.1% formic acid and B: 90:10 IPA:ACN+10mM ammonium formate+0.1% formic acid. The column used was an Acquity Premier BEH C18 1.7 μm, 2.1 x 50mm with an injection volume of 1 μL. Samples were injected using Agilent 6530 QTOF collecting data from 120m/z to 1200m/z at an acquisition rate of 2 spectra/sec.

#### Negative ionization mode

Mobile phase consists of A: 60:40 ACN:H_2_O+10mM ammonium acetate and B: 90:10 IPA:ACN+10mM ammonium acetate. The column used was an Acquity Premier BEH C18 1.7 μm, 2.1 x 50mm with an injection volume of 5 μL. Samples were injected using Agilent 6550 QTOF collecting data from 60m/z to 1200m/z at an acquisition rate of 2 spectra/sec.

#### Data Processing

The general workflow for data processing was completed using MS-DIAL (15), followed by data cleanup using MS-FLO (16). In brief, raw files were first converted via Abf Converter and processed using MS-DIAL. Once the results were exported from MS-DIAL, a blank reduction was done based on the max peak height relative to blank average height. Using MS-FLO, potential duplicates and isotopes were checked and deleted, if identified. Next, MS/MS spectra were assessed before combining adducts. Peaks were annotated in manual comparison of MS/MS spectra and accurate masses of the precursor ion to spectra given in the Fiehn laboratory’s LipidBlast spectral library (17). Additional peaks were found by manual curation of sample chromatograms on a scan-by-scan basis. MassHunter Quant software was then used to verify peak candidates based on peak shape, peak height reproducibility, and retention time reproducibility in replicate samples. Valid and reproducible peaks were confirmed with MS/MS spectra with the aim of increasing overall peak annotations in both positive and negative modes. These manually curated compounds were incorporated into a .txt file that had a list of accurate masses and retention times for the lipidomics platform. All data were normalized to the sum of internal standards and are reported as peak heights for quantification ion at a specific retention time. Peak heights are used for two reasons: 1) they are more precise for low-abundant metabolites than peak areas, due to the larger influence of baseline determinations on areas compared to peak heights, and 2) co-eluting ions or peaks are harder to deconvolute on peak areas than on peak heights.

### Biogenic Amines and Small Molecule Metabolites

After dry down, samples were reconstituted in 100 μL of 80:20 ACN:H_2_O + internal standard mixture. The samples were then vortexed for 10s, sonicated for 5 min, and centrifuged for 2 min at 16,000 x *g*. 45 μL was transferred to two separate glass amber vials with micro inserts for positive and negative ionization modes. All data were normalized to the sum of internal standards.

#### Positive ionization mode

Mobile phase consists of A: 100% H_2_O + 10mM ammonium formate + 0.125% formic acid and B: 95:5 ACN:H_2_O + 10 mM ammonium formate + 0.125% formic acid. The column used was an Acquity Premier UPLC BEH Amide 1.7 μm, 2.1 x 50mm with an injection volume of 1 μL. Samples were injected into an Agilent 1290 UHPLC/Sciex TripleTOF 6600 mass spectrometer.

#### Negative ionization mode

Mobile phase consists of A: 100% H_2_O + 10mM ammonium formate + 0.125% formic acid and B: 95:5 ACN:H_2_O + 10 mM ammonium formate + 0.125% formic acid. The column used was an Acquity Premier UPLC BEH Amide 1.7 μm, 2.1 x 50mm with an injection volume of 1 μL. Samples were injected into an Agilent 1290 UHPLC/Sciex TripleTOF 6600 mass spectrometer.

#### Data Processing

Chromatograms first underwent a quality control check in which internal standards were examined for consistency of peak height and retention time. Raw data files were then processed using an updated version of MS-DIAL software (15), which identified and aligned peaks and then annotated those peaks using both an in-house mzRT library and MS/MS spectral matching with NIST/MoNA libraries. All MS/MS annotations were then manually curated to ensure that only high-quality compound identifications were included in the final analysis.

### Primary Metabolites

Samples are shaken at 30°C for 1.5 h. Then, 91 μL of N-trimethylsilyl-N-methyl trifluoroacetamide (MSTFA) + fatty acid methyl esters (FAMEs) internal standards are added to each sample and are shaken at 37°C for 0.5 h to finish derivatization. Samples are then vialed, capped, and injected onto the 7890A GC coupled with a LECO TOF. Derivatized sample volume is 0.5 uL using a split-less method onto a RESTEK RTX-5SIL MS column with an Integra-Guard at 275C with a helium flow rate of 1 mL/min. The GC oven was set to hold at 50°C for 1 min, then ramp to 20°C/min to 330°C and then hold for 5 min. The transfer line was set to 280°C while the EI ion source was set to 250°C. The mass spectrometer parameters have been published elsewhere (18) and collect data from 85m/z to 500m/z at an acquisition rate of 17 spectra/sec.

#### Data Processing

ChromaTOF 4.72 was used for data preprocessing without smoothing, 3 s peak width, baseline subtraction just above noise level, and automatic mass spectral deconvolution and peak detection at signal/noise levels of 5:1 throughout the chromatogram. Apex masses were reported for use in the BinBase algorithm. Result *.txt files were then exported to a data server with absolute spectra intensities and further processed by a filtering algorithm implemented in the metabolomics BinBase database. The BinBase algorithm (rtx5) used the settings: validity of chromatogram (<10 peaks with intensity >10^7 counts s-1), unbiased retention index marker detection (MS similarity>800, validity of intensity range for high m/z marker ions), retention index calculation by 5th order polynomial regression. Spectra were cut to 5% base peak abundance and matched to database entries from most to least abundant spectra using the following matching filters: retention index window ±2,000 units (equivalent to about ±2 s retention time), validation of unique ions and apex masses (unique ion must be included in apexing masses and present at >3% of base peak abundance), mass spectrum similarity must fit criteria dependent on peak purity and signal/noise ratios and a final isomer filter. Failed spectra are automatically entered as new database entries if s/n >25, purity <1.0, and presence in the biological study design class was >80%. All thresholds reflect settings for ChromaTOF 2.32. Quantification is reported as peak height for reasons noted above.

### Bioinformatics & Statistical Analysis

All data processing and statistical analyses used R v4.4.1 (19) and Bioconductor v3.19 (20). Abundance values were read in using either readxl v1.4.3 (21) or readr v2.1.5 (22), with feature id and sample id cleaning using janitor v2.2.0 (23) and dplyr v1.1.4 (24). Plots were generated using either singly or in combination packages: ggplot2 v3.5.1 (25); geom_sina from ggforce v0.4.2 (26); patchwork v1.3.0 (27); ComplexHeatmap v2.20.0 (28). Analysis workflows were coordinated using targets v1.8.0 (29).

All four omics datasets (transcriptomics, biogenic amines, primary metabolism, lipidomics) were processed and analyzed separately as they were acquired in separate runs of instrumentation. Omics features were kept for further analysis if they were present (raw abundance >= 10) in 75% or more in either the tumor or adjacent nontumor samples (visualizationQualityControl v0.5.1) (30). All data sets were stored in R and manipulated as DESeq2 datasets (v1.44.0) (31). Sample normalization used estimateSizeFactors from DESeq2.

#### Quality Control (QC)

Sample - sample correlations were calculated using information-content-informed Kendall-tau (ICI-Kt, ICIKendallTau v1.2.2) (32). Outlier samples to be removed prior to differential analysis were based on scores calculated from the median ICI-Kt values within tumor and adjacent nontumor treatments and outliers detected using boxplot.stats. The outliers detected and final sample numbers across all analyses can be found in **Supplementary Figure 2** and **Supplemental Table 1**, respectively.

#### Principal Component Analysis (PCA)

Missing values were set to zero, and abundances were log-transformed using log(x + 1) (log1p function in R), and principal components calculated using the prcomp function, with centering and no scaling. Principal component variances and percentages of total were calculated using visqc_score_contributions.

#### Differential Comparisons

Patients that had either a tumor or adjacent nontumor sample removed from outlier detection had the other sample removed prior to differential analysis. Any duplicate samples were collapsed using the collapseReplicates function from DESeq2. Statistics were calculated using DESeq2, with dispersions fit to the mean intensity using the paremetric method for transcriptomics, and the local method for the metabolomics datasets. The linear model for the differential analysis included the patient to account for the paired nature of the samples, specifically using the design of ∼ patient + treatment, and then extracting the contrast of treatment in the DESeq results. P-values were adjusted using the Benjamini-Hochberg method (33).

#### Annotation Enrichment

Reactome pathway annotations for Ensembl genes were downloaded from the reactome pathway website on 2024-02-23 (34). Reactome pathway annotations for ChEBI metabolites were downloaded from the reactome pathway website. Metabolites were mapped to ChEBI by a combination of InChiKey’s generated by OpenBabel (v3.1.1) (35) from the download of ChEBI to InChI, as well as through mapping of KEGG compounds. Lipids were annotated with their lipid class, chain lengths, total chain lengths, and degree of unsaturation using custom code to parse the lipid IDs provided by West Coast Metabolomics.

Hypergeometric enrichment used all annotated features as the universe, and those features with an Padj ≤0.01 as the significant set, with features split by log-fold-change direction (positively and negatively changed), using the hypergeometric_feature_enrichment in categoryCompare2 v0.200.2 (36). Enrichment p-values were adjusted using the Benjamini-Hochberg method. Binomial enrichment used all features regardless of p-value, using the binomial_feature_enrichment function in categoryCompare2, with annotations that have at least six features, the null hypothesis that the ratio of positively to negatively changed features should be 0.5, and adjusting p-values using the Benjamini-Hochberg method. Lipid annotations tested included the lipid class, individual chain length, total chain length, degree of unsaturation, as well as various combinations of the annotations.

#### Spearman Correlation of Transcripts and Metabolites

Correlations of all metabolites were calculated with all of the transcripts across the samples common to all omics methods, using Spearman correlation, with zero representing a missing value. Significantly changed transcripts (p-adjusted ≤0.01) with significant correlations (p-adjusted ≤0.01) to either compounds (biogenic amines & primary metabolism) or lipids that are associated with a significant binomial enriched category and with log-fold-change in the same direction as the overall direction of the enrichment (pathway or lipid annotation, p-adjusted ≤0.05) were extracted. The Spearman correlation was used as input for calculating the Euclidian distances between the metabolites and the genes, and then clustered using hierarchical clustering via hclust, and ordered using dendsort v0.3.4 (37). Clusters of metabolites and transcripts were defined in two passes. The first was by splitting the hierarchical clustering into x clusters using cutree, defined by the number of clusters desired. Subsequently, chosen clusters were split into sub-clusters by pruning away branches not in the desired cluster (via the prune function of dendextend v1.18.0 (38)), and then splitting the remainder again into the number of desired clusters using cutree.

Hypergeometric enrichment of transcripts and metabolites in each cluster used all measured features as the universe, and the members of the cluster as the significant set. P-values were adjusted using the Benjamini-Hochberg method.

## RESULTS

### Patient Characteristics

Eight human HCC tumors and adjacent paired non-tumor tissue were collected and used for analysis. All patients were non-Hispanic and consisted of four male and four female patients with a mean age of 67.3 years. Mean body mass index (BMI) was 31.7 kg/m^2^, with seven of the eight patients having a BMI greater than MASLD cardiometabolic criteria of 25 kg/m^2^. Five patients were diabetic or taking medications for diabetes, and two were hyperlipidemic or taking lipid-lowering therapies. Combined, seven of the eight patients met at least one cardiometabolic MASLD criteria, while one patient met criteria for MetALD due to a history of prior alcohol use disorder (∼140g of alcohol/day) (11). None of the patients had documented antibodies for hepatitis B or C. Patient characteristics and available clinical data are summarized in **Figure 1A**.

**Figure 1.**
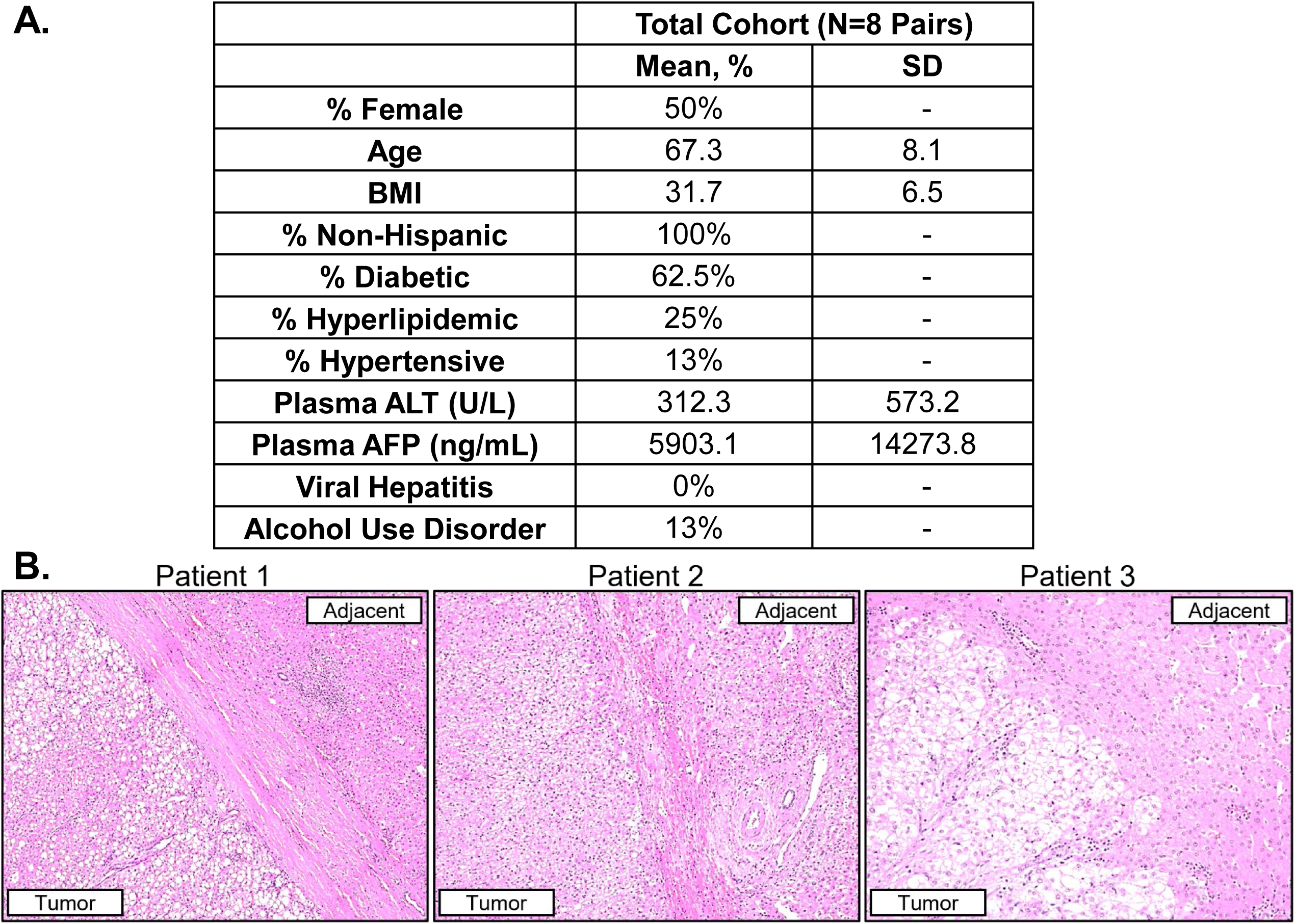
Patient Characteristics and Tumor Histology. (**A**) Patient cohort demographics and clinical data. (**B**) H&E stained liver sections from 3 patients highlighting tumor and adjacent nontumor tissue used in the analysis.

Hematoxylin and eosin-stained images of tumor and adjacent nontumor tissue were verified by an independent pathologist. Tumor tissues were enriched in lipid vacuoles with clear inflammatory foci present in both the tumor and nontumor tissue (**Fig. 1B**). Another histological feature was the presence of a fibrotic capsule surrounding the tumor tissue, which is commonly observed in HCC (39). Taken together, this patient cohort resembles that of the metabolically-driven steatohepatitic variant of HCC.

### Complement Cascade and Scavenging Pathways Are Uniquely Repressed in HCC

To characterize the tumor transcriptome, we conducted bulk RNA-sequencing on paired tumor and adjacent nontumor tissue. We identified 1,679 genes that met a pre-determined statistical cutoff of (p-adjusted value [Padj] ≤0.01). Of the 1679 genes identified, 1398 (83.3%) were reduced while 281 (16.7%) were increased in tumor versus nontumor tissue across paired samples. A list of all differentially expressed genes can be found in **Supplemental Table 2**. Principal component analysis (PCA) revealed the transcriptome of tumors were profoundly different from that of adjacent nontumor tissue, with PC1 accounting for 28% and PC2 accounting for 12% of the variance (**Fig. 2A**). Genes encoding proteins involved in metal chelation and scavenging of free radicals were largely repressed in tumor tissue (*MT1F, MT1X, MT2A;* **Fig. 2B, C**). Consistent with a previous report (40), we also identified significant reductions in the 2-hydroxy acid oxidases (*HAO1*: Log2-FC=-2.31; Padj=1.64E-5; *HAO2*: Log2-FC=-5.33; Padj=6.21E-20) in tumor tissue (**Fig. 2B, C**). Another gene of interest was that of solute carrier family 25 member 47 (*SLC25A47*), which encodes a liver-specific mitochondrial NAD+ transporter that, when deleted, leads to impaired mitochondrial fatty acid oxidation, liver fibrosis, and HCC tumorigenesis in mice (41, 42). Consistent with its role in chronic liver disease, *SLC25A47* was significantly repressed in HCC (Log2FC=-5.20; Padj=2.63E-10). Further, genes involved in complement activation (*C9, C1QTNF1*) and lipoprotein metabolism (*APOF*) were also reduced in HCC tumor tissue across paired samples (**Fig. 2B-D**).

**Figure 2.**
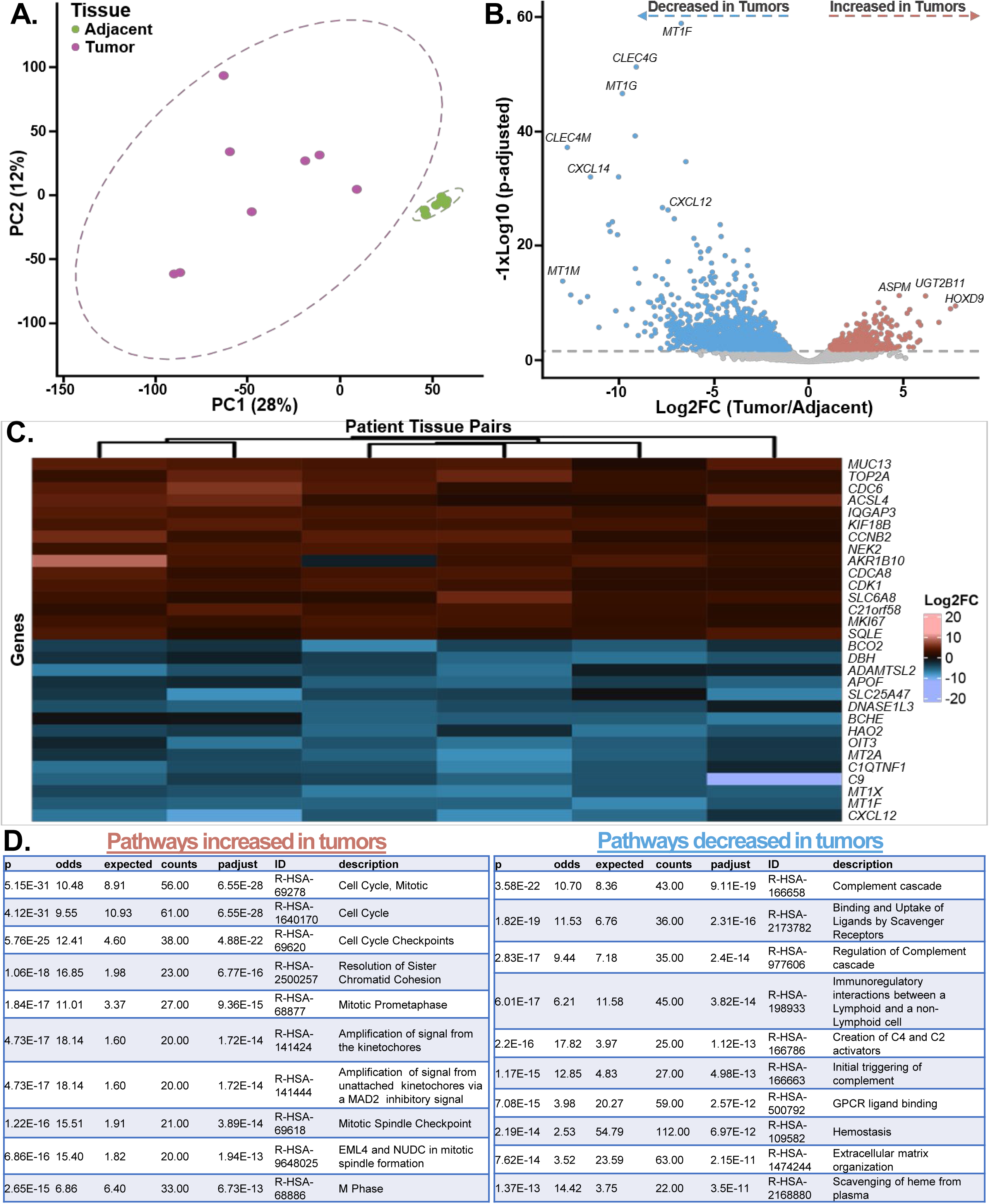
Transcriptomic Profiling of Tumor and Adjacent Nontumor Tissue. (**A**) Principal component analysis of tumor (purple) and adjacent nontumor (green) tissue. (**B**) Volcano plot highlighting genes which increased (in red) or decreased (in blue) in tumors relative to nontumor tissue. (**C**) A heatmap highlighting the top 15 increased (red/pink) and top 15 decreased (blue/purple) genes in tumor relative to nontumor tissue. (**D**) Reactome pathway enrichment analyses of RNA-sequencing data. The top 10 pathways increased in tumors (left; red) and decreased in tumors (right; blue) relative to nontumor tissue.

We then assessed gene transcripts increased in HCC relative to nontumor tissue. This analysis identified genes commonly associated with cellular proliferation pathways (**Fig. 2D**), including increased DNA topoisomerase II alpha (*TOP2A*: Log2FC=3.79; Padj=5.44E-9), cell division cycle 6 (*CDC6*: Log2FC=3.76; Padj=3.51E-7), and marker of proliferation Ki-67 (*MKI67*: Log2FC=2.93; Padj=5.93E-9; **Fig. 2B, C**). Other noteworthy genes elevated in human HCC were related to fatty acid and steroid metabolism, including acyl-CoA synthetase long chain family member 4 (*ACSL4*: Log2FC=3.72; Padj=1.17E-6), squalene epoxidase (*SQLE*: Log2FC=2.90; Padj=1.32E-7), and hydroxysteroid 17-beta dehydrogenase 7 (*HSD17B7*: Log2FC=1.35; Padj=3.34E-3; **Fig. 2B, C**). Taken together, pathways involved in cell cycle control were largely increased with Padj ranging from 6.73E-13 for R-HSA-68886 “M Phase” to 6.55E-28 for R-HSA-1640170 “Cell Cycle” (**Fig. 2D**; **Supplemental Table 3**). Genes involved in complement signaling, scavenging receptors, and extracellular matrix organization were decreased in tumor tissue, specifically R-HSA-166658 “Complement cascade” with a Padj of 9.11E-19, R-HSA-2173782 “Binding and Uptake of Ligands by Scavenger Receptors” with Padj of 2.31E-16, and R-HSA-1474244 “Extracellular matrix organization” with Padj of 2.15E-11 (**Fig. 2D; Supplemental Table 4**).

### Phospholipid and Triacylglycerol Saturation is Increased in HCC

Given the profound lipid vacuole accumulation observed by histology in **Fig. 1B**, we next utilized LC-HRMS/MS to determine which lipid species were altered in tumor versus adjacent nontumor tissue. PCA revealed the lipidome of tumors were profoundly different from that of adjacent nontumor tissue, with PC1 accounting for 32% and PC2 accounting for 12% of the variance (**Fig. 3A**). We identified 292 lipid species that met a pre-determined statistical cutoff of Padj ≤0.01. Of the 292 lipids significantly changes, a majority were unannotated (250, or 86%) while 42 were previously annotated lipids (14%; **Supplemental Table 5**). The 15 most increased annotated lipids in HCC relative to nontumor were saturated and monounsaturated phospholipids, including PG16:0_16:0 (Log2FC=2.03; Padj=2.52E-4), PC35:0 (Log2FC=1.97; Padj=2.25E-4), PC16:0_14:0 (Log2FC=1.91; Padj=5.66E-6), and PC14:0_16:1 (Log2FC=1.47; Padj=8.01E-3; **Fig. 3B, C**; **Supplemental Table 5**). Meanwhile, the 15 most repressed annotated lipids in tumors were polyunsaturated phospholipids (PG18:1_18:2; Log2FC=-3.36; Padj=1.27E-6) and triglycerides (TG54:6; Log2FC=-2.52; Padj=6.27E-6), ceramides (CER16 lactosyl-ceramide d18:1_16:0; Log2FC=-1.66; Padj=5.61E-3), and acylcarnitines (CAR18:2; Log2FC=-1.85; Padj=4.73E-4; **Fig. 3B, C**; **Supplemental Table 5**), relative to nontumor tissue.

**Figure 3.**
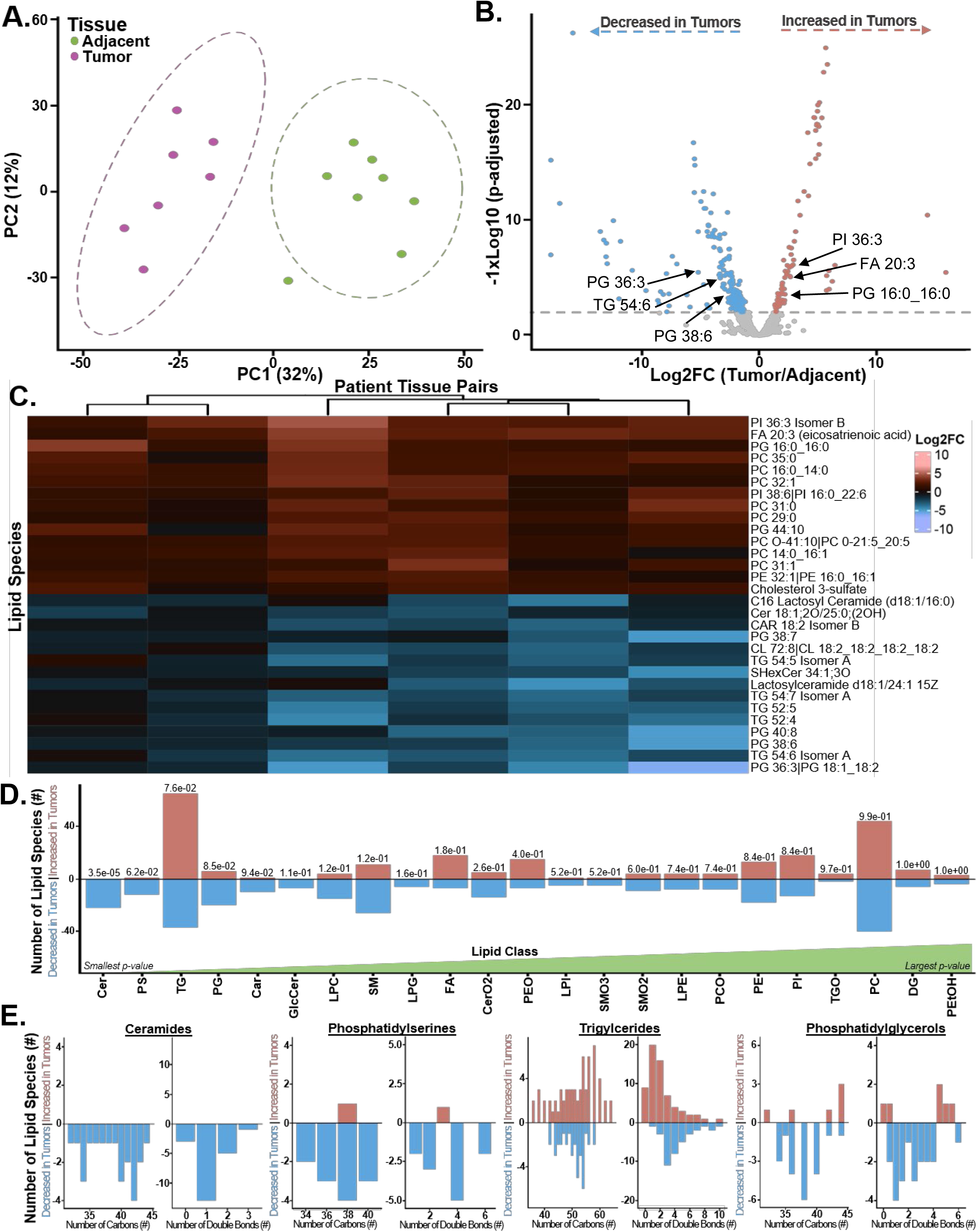
Lipidomic Profiling of Tumor and Adjacent Nontumor Tissue. (**A**) Principal component analysis of tumor (purple) and adjacent nontumor (green) tissue. (**B**) Volcano plot highlighting lipids which increased (in red) or decreased (in blue) in tumors relative to nontumor tissue. (**C**) A heatmap highlighting the top 15 increased (red/pink) and top 15 decreased (blue/purple) lipids in tumor relative to nontumor tissue. (**D**) Binomial analysis of lipid enrichment across 22 lipid classes (x-axis). The y-axis denotes the total number (#) of lipid species increased (red) or decreased (blue) in tumors relative to nontumor tissue. Padj values increase from left to right across lipid classes. (**E**) The number of carbons (left) and number of double bonds (right) in each of the top 4 lipid classes initially identified by binomial analysis in Fig. 3D (ceramides, phosphatidylserines, triglycerides, and phosphatidylglycerols).

We next performed binomial enrichment analysis of the lipids to determine which lipid classes and other lipid groups were most differentially affected across tumor and adjacent nontumor tissue (**Supplemental Table 6**). Of the 23 lipid classes, ceramides were the only lipid class significantly different and decreased in HCC (Padj=3.5E-5). Of importance to note, all 22 ceramide lipids detected by LC-HRMS/MS, regardless of the number of carbons or degree of saturation, were decreased in tumors (**Fig. 3D, E**). Similar to ceramides, phosphatidylserines (Padj=6.2E-2), phosphatidylglycerols (Padj=8.5E-2), and acylcarnitines (Padj=9.4E-2) all trended to decrease in HCC. On the contrary, triglycerides (Padj=7.6E-2) and non-esterified fatty acids (NEFAs; Padj=1.8E-1) tended to increase in tumors relative to nontumor tissue. Of the total triglyceride pool, 100% of saturated triglycerides (Padj=6.3E-2; class_total_db:TG:0 in **Supplemental Table 6**), 95% of MUFA-enriched triglycerides (Padj=1.0E-3; class_total_db:TG:1 in **Supplemental Table 6**), and 92% of triglycerides with ≤ two double bonds were significantly increased in tumors relative to nontumor tissue. We observed similar increases in fatty acyl chain saturation in phosphatidylcholine (**Supplementary Figure 3A**) and in NEFAs (**Supplementary Figure 3B**) of tumor tissue, while their respective unsaturated counterparts tend to be decreased in HCC (**Fig. 3E; Supplementary Figure 3A, B**). Taken together, and relative to adjacent nontumor tissue, human HCC exhibits a lipid signature that consists of: 1) statistically significant reductions in ceramides, 2) statistically significant accumulation of saturated- and monounsaturated NEFAs, triglycerides, and phosphatidylcholine, and 3) plausible reductions in phosphatidylserine lipids.

### Identification of Novel Gene-Lipid Associations in Human HCC

Utilizing the bulk RNA-sequencing and lipidomics platforms, we next sought to identify novel gene-lipid associations across paired tumor and adjacent nontumor tissue. Utilizing only significantly altered gene transcripts from bulk RNA-sequencing in **Fig. 2** and lipids associated with significantly enriched lipid categories, we employed Spearman correlation analyses between gene transcripts and lipid species. We initially identified a total of 5941 gene-lipid associations (**Fig. 4A; Supplementary Figure 4; Supplementary Table 7**). When applied to the heatmap matrix, core enrichment was observed across 4 gene (y-axis) and 3 lipidomic (x-axis) subclusters (**Fig. 4A; Supplementary Figure 4**). Utilizing hypergeometric enrichment, gene cluster 1 was enriched in extracellular matrix genes which associated with the lipidomic cluster (lipidomic cluster 2) enriched in ceramide lipid species (**Fig. 4A; Supplemental Table 8**). No other gene-lipid clusters yielded significant enrichments of gene or lipid annotations.

**Figure 4.**
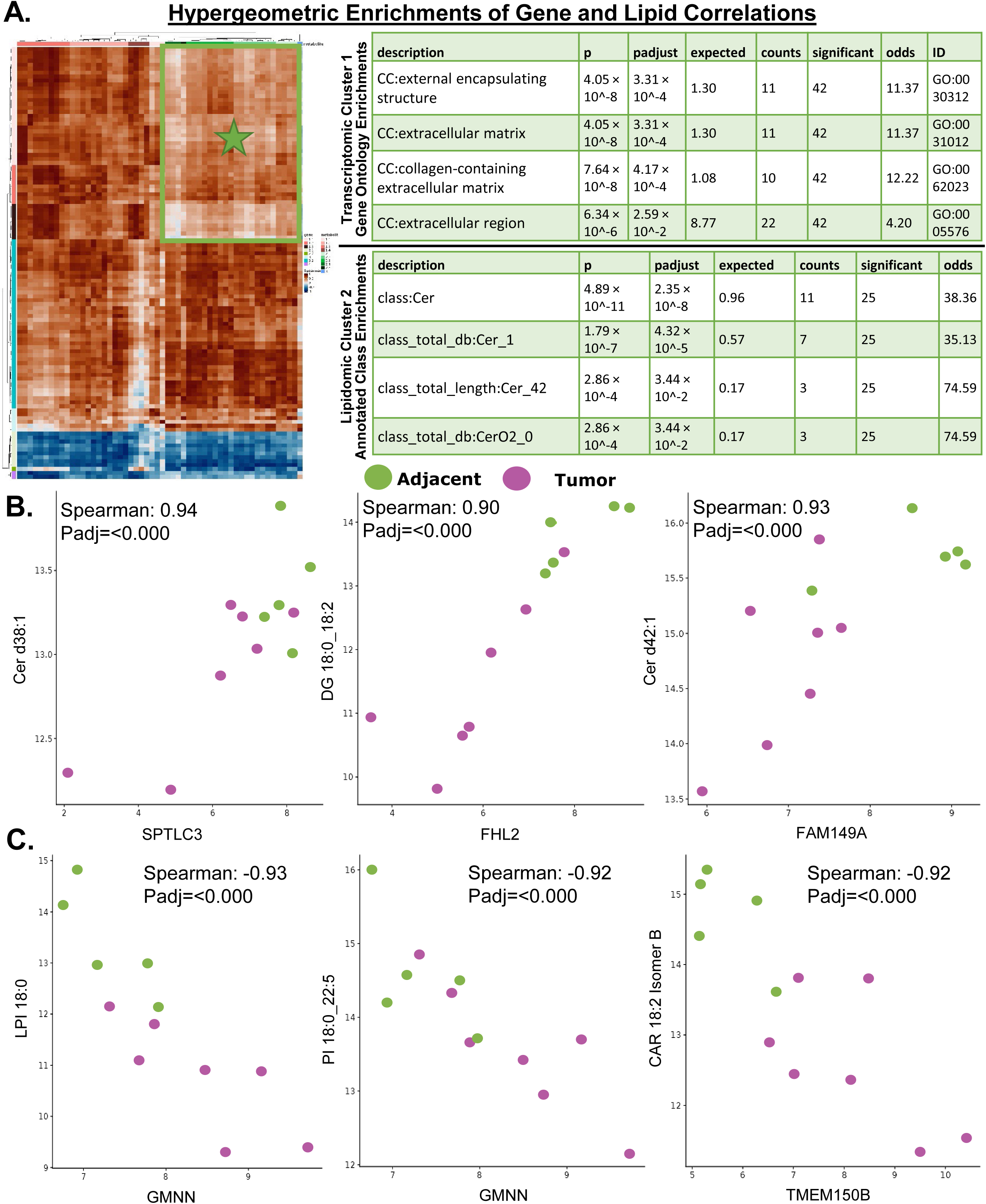
Gene-Lipid Correlations in HCC. (**A**) A heatmap highlighting all 5941 gene-lipid associations and hypergeometric enrichments of gene-lipid correlations across heatmap subclusters. X-axis=lipid clustering (i.e. metabolite); Y-axis=gene clustering. Spearman correlation coefficient ranging from -1 (blue) to +1 (brown). The table represent gene ontology enrichment of transcriptomic cluster 1 and its association with lipidomic cluster 2, identified by the green star in the heatmap. (**B, C**) Three highly significant associations in the positive (**B**) and negative (**C**) direction between specific transcripts and lipid species across tumor (purple) and adjacent nontumor tissue (green).

Then, of the initial 5941 associations, we found 186 meeting both the predetermined statistical cutoff of Padj<0.01 and a Spearman’s rank correlation coefficient of│0.90│. Of the 186, 17 were negatively associated while 169 were positively associated with specific lipid species (see complete table in **Supplemental Table 7**). For instance, serine palmitoyltransferase long chain base subunit 3 (*SPTLC3*) positively associated with ceramides d38:1 (Spearman: 0.94), ceramide d39:1 (Spearman: 0.91), ceramide d41:2 (Spearman: 0.90), and ceramide d43:1 (Spearman: 0.95), with tumors exhibiting lower levels of both gene and lipid levels (**Fig. 4B; Supplemental Table 7**). Further, the family with sequence similarity 149, member A gene (*FAM149A*) also positively associated with ceramides 18:0;2O_24:0 (Spearman 0.93; **Fig. 4B**) and ceramide d42:1 (Spearman: 0.93; **Supplemental Table 7**). Other gene-lipid associations revealed a positive association of four and a half LIM domain protein 2 (*FHL2*) with DG18:0_18:2 (Spearman: 0.90; **Fig. 4B**) and PE34:2 (Spearman: 0.91; **Supplemental Table 7**) levels across tumor and adjacent nontumor tissue. Of the negative associations, the geminin DNA replication inhibitor (*GMNN*) gene was increased in tumors which negatively associated with LPI:18:0 (Spearman: -0.93) and PI18:0_22:5 (Spearman: -0.92) lipids (**Fig. 4C**). Meanwhile, a key autophagic regulatory gene (43), transmembrane protein 150B (*TMEM150B*; otherwise known as *DRAM-3*), is increased in tumors and negatively associated with acylcarnitine CAR18:2 (Spearman: - 0.92; **Fig. 4C**). Taken together, this work identified novel gene-lipid relationships in human HCC that warrant mechanistic validation in preclinical cancer models.

### Amino Acid and Fatty Acid Transport and Metabolism Pathways Are Repressed in HCC

We next sought to ask whether specific metabolites were uniquely altered across paired tumor and adjacent nontumor tissue, and how these changes might associate with transcript levels. We performed metabolomics utilizing two MS-based platforms by West Coast Metabolomics. Using the primary metabolism platform (**Figs. 5A-C**), we identified a total of 47 metabolites that met a predetermined statistical cutoff of Padj<0.01. Of the 47, only 9 (19.1%) have been previously annotated (**Supplemental Table 9-PM in column Q**). Consistent with the low number of statistically significant metabolites, PCA analysis revealed an overlap between tumor and adjacent nontumor tissue with PC1 accounting for 21% and PC2 accounting 11% of the total variance across groups (**Fig. 5A**). The top 3 most increased metabolites in tumors relative to nontumor were tartaric acid (Log2FC=2.68; Padj=1.28E-13), serotonin (Log2FC=1.17; Padj=6.00E-03), and phosphate (Log2FC=0.87; Padj=7.50E-03). The top 3 most abundant metabolites in adjacent nontumor tissue were cysteine (Log2FC=-1.99; Padj=7.50E-03), xanthosine (Log2FC=-1.90; Padj=1.30E-03), and cellobiose (Log2FC=-1.36; Padj=9.80E-03).

**Figure 5.**
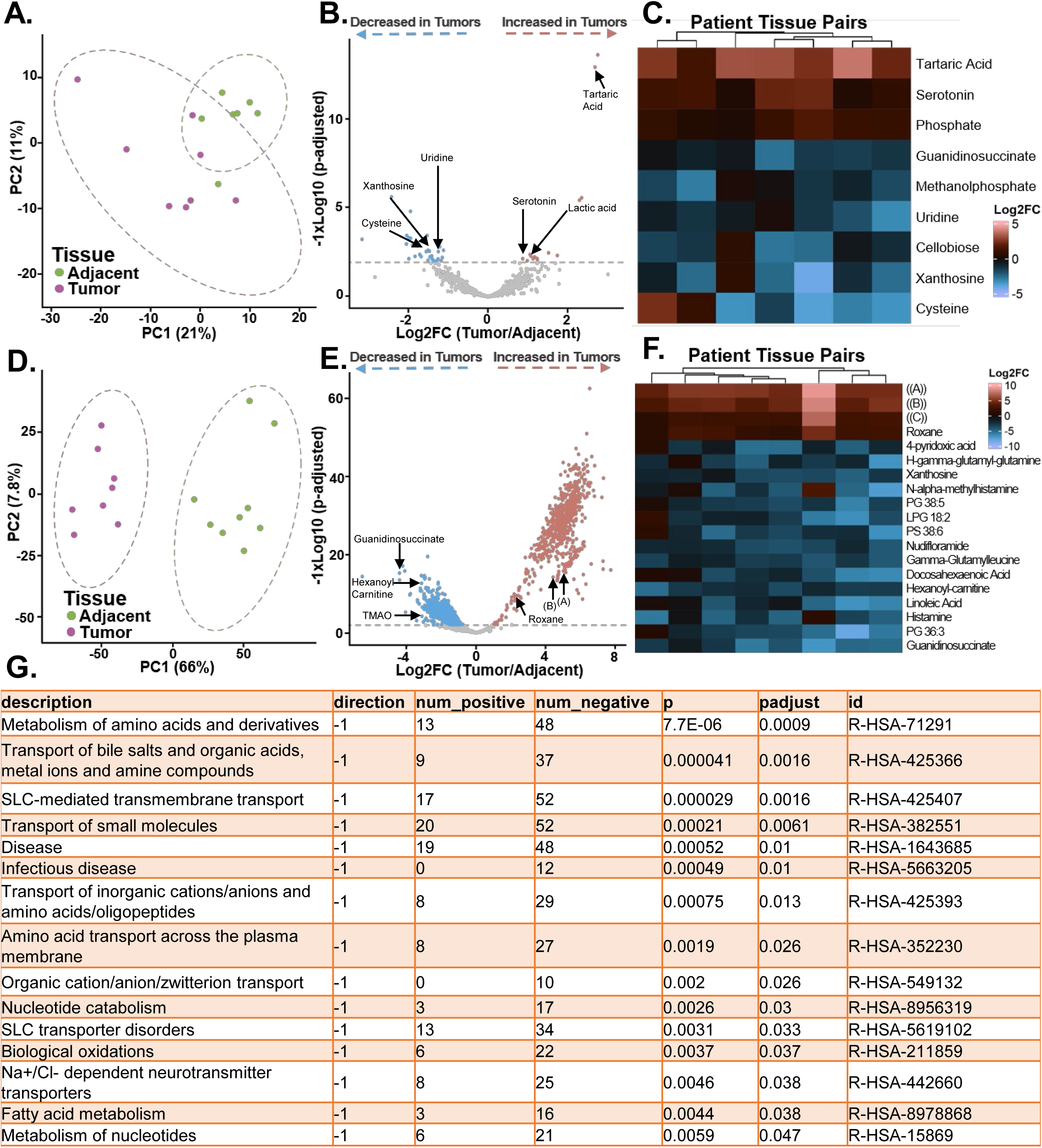
Metabolomic Profiling of Tumor and Adjacent Nontumor Tissue. Metabolomics was performed on tumor and adjacent nontumor tissue utilizing the primary metabolism (**A**-**C**) and biogenic amine (**D**-**F**) platforms by West Coast Metabolomics. (**A, D**) Principal component analysis from tumor (purple) and adjacent nontumor (green) tissue from metabolomic platforms. (**B, E**) Volcano plot highlighting metabolites which increased (in red) or decreased (in blue) in tumors relative to nontumor tissue. (**C, F**) Heatmaps highlighting the most increased (red/pink) and decreased (blue/purple) metabolites in tumor relative to nontumor tissue. (**G**) Reactome pathway enrichment analyses highlighting the most significantly altered pathways across metabolomic datasets.

Using the biogenic amine platform (**Figs. 5D-E**), we identified a total of 1,649 significantly altered (Padj<0.01) metabolites with 317 (19.2%) of those metabolites previously annotated (**Supplemental Table 9-bioamines in column Q**). Given the significant number of altered metabolites, PCA analysis revealed biogenic amine profiles were profoundly different in tumors, with PC1 accounting for 66% and PC2 accounting for 7.8% of the variance (**Fig. 5D**). The top 4 most increased metabolites in tumors relative to nontumor were benzyldimethyltetradecylammonium cation ([A] in **Figs. 5E-F**; Log2FC=5.17; Padj=4.16E-16), N-benzyl-N,N-dimethyl-1-hexadecanaminium cation ([B] in **Figs. 5E-F**; Log2FC=4.43; Padj=6.84E-15), benzyldimethylstearylammonium cation ([C] in **Figs. 5E-F**; Log2FC=2.61; Padj=1.13E-07), and roxane (Log2FC=2.41; Padj=3.16E-10). The most decreased metabolites in tumors relative to adjacent nontumor tissue included guanidinosuccinate (Log2FC=-4.26; Padj=8.83E-18), histamine (Log2FC=-3.40; Padj=4.52E-9), hexanoyl-carnitine (Log2FC=-3.03; Padj=5.91E-15), and docosahexaenoic acid (Log2FC=-3.01; Padj=1.06E-06). Combining the two platforms together, we then asked which pathways were enriched across these metabolites. Notably, pathways related to amino acid metabolism and transport (R-HSA-71291; Padj=0.0009), transport of bile salts (R-HSA-425366; Padj=0.0016) and small molecules (R-HSA-382551; Padj=0.0061), and nucleotide (R-HSA-8956319; Padj=0.03) and fatty acid metabolism (R-HSA-8978868; Padj=0.038) were all decreased in HCC tumors relative to nontumor tissue (**Fig. 5G; Supplemental Table 10**).

### Identification of Novel Gene-Metabolite Associations in Human HCC

Utilizing the bulk RNA-sequencing and combined metabolomic platforms, we next sought to identify novel gene-metabolite associations across paired tumor and adjacent nontumor tissue. Utilizing significantly altered gene transcripts from bulk RNA-sequencing in **Fig. 2**, we employed Spearman correlation analyses across both metabolomic platforms. We identified a total of 2800 gene-metabolite associations (**Fig. 6A; Supplementary Figure 5; Supplementary Table 11**). When applied to the heatmap matrix, core enrichment was observed across 4 gene (y-axis) and 5 metabolite (x-axis) subclusters (**Fig. 6A; Supplementary Figure 5**). Utilizing enrichment analyses across these subclusters, gene clusters 1 and 2 were both associated with the metabolomic cluster 1 linking fatty acid oxidation and extracellular matrix gene transcripts to alterations in nucleotide repair pathways and Akt signaling (**Fig. 6A; Supplementary Figure 6**; **Supplemental Table 12**).

**Figure 6.**
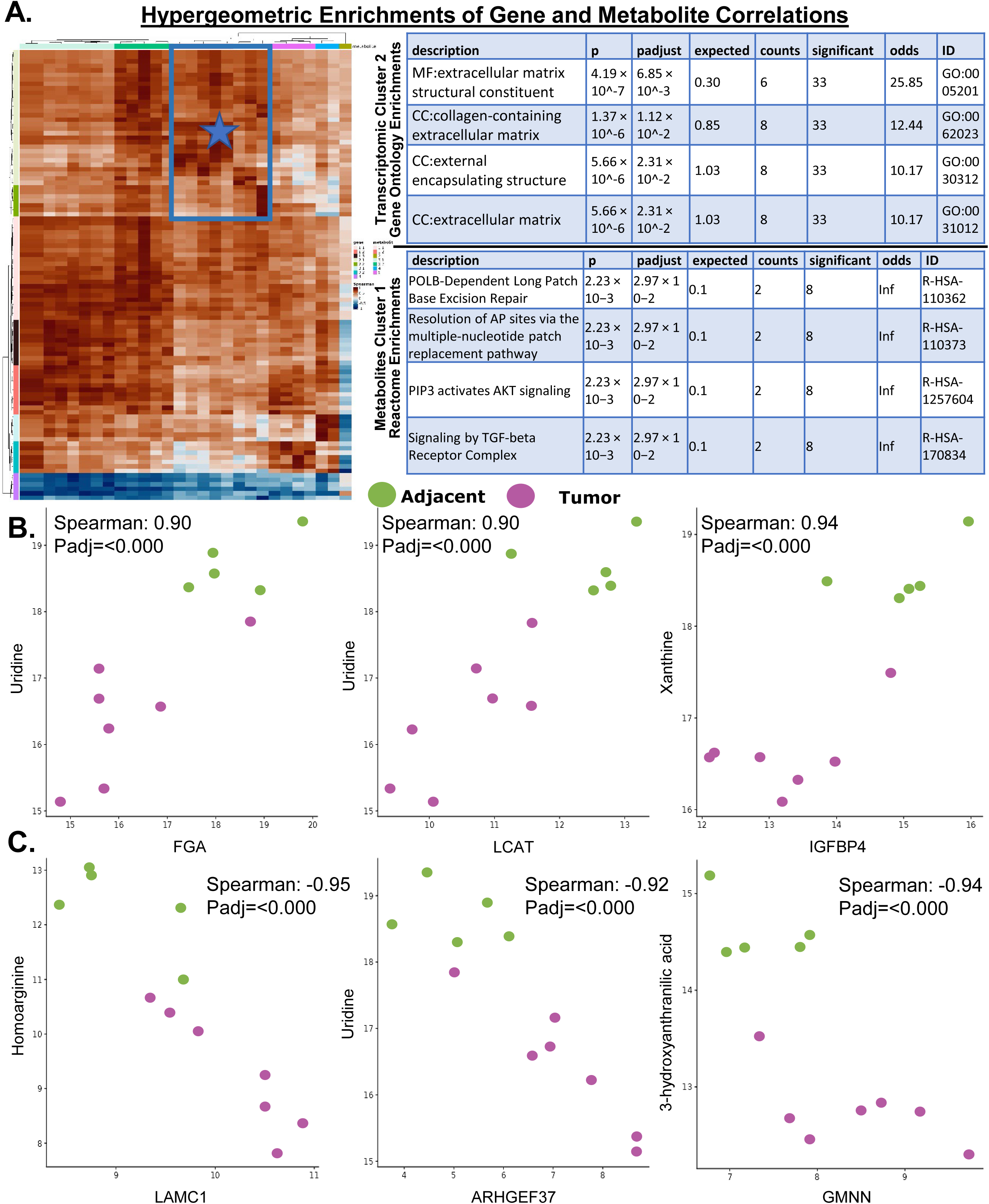
Gene-Metabolite Correlations in HCC. (**A**) A heatmap highlighting all 2800 gene-metabolite associations and hypergeometric enrichments of gene-metabolite correlations across heatmap subclusters. X-axis=metabolite clustering (i.e. biogenic amines + primary metabolites); Y-axis=gene clustering. Spearman correlation coefficient ranging from -1 (blue) to +1 (brown). The table represents gene ontology enrichment of transcriptomic cluster 2 and its association with Reactome enrichment of metabolomic cluster 1, identified by the blue star in the heatmap. (**B, C**) Three highly significant associations in the positive (**B**) and negative (**C**) direction between specific transcripts and metabolites across tumor (purple) and adjacent nontumor tissue (green).

Of the initial 2800 associations, we found 117 who met both the predetermined statistical cutoff of Padj<0.01 and a Spearman’s rank correlation coefficient of│0.90│. Of the 117, 9 were negatively associated while 108 were positively associated with specific annotated metabolites (see complete table in **Supplemental Table 11**). Consistent with reports that uridine inhibits HCC (44), we observed reductions in uridine levels in tumors relative to nontumor tissue (**Fig. 6B, C**). Notably, uridine levels positively associated with transcripts involved in coagulation (factor fibrinogen alpha chain; *FGA*; Spearman: 0.90), cholesterol esterification (lecithin-cholesterol acyltransferase; *LCAT*; Spearman: 0.90), and demethylation of C4-methylsterols (45) (fatty acid hydroxylase containing 2; *FAXDC2*; Spearman: 0.92). Meanwhile, uridine only negatively associated with one gene, the Rho Guanine Nucleotide Exchange Factor 37 (*ARHGEF37*; Spearman: -0.92; **Fig. 6B-C; Supplemental Table 11**). Aside from uridine, we observed reductions in xanthine levels in HCC tumors that positively associated with insulin like growth factor binding protein 4 (*IGFBP4*; Spearman: 0.94; **Fig. 6B**), reticulophagy regulator 1 (*RETREG1*; Spearman: 0.91), and negatively associated with *GMNN* (Spearman: -0.94; **Supplemental Table 11**). Other notable negative associations include laminin subunit gamma 1 (*LAMC1*) gene with homoarginine (Spearman: -0.95) and the tryptophan metabolite, 3-hydroxyanthranilic acid (3-HAA), with *GMNN* (Spearman:-0.94; **Fig. 6C**).

## DISCUSSION

In this manuscript, we aimed to determine how the metabolome, lipidome, and transcriptome were differentially affected in human steatohepatitic tumors versus paired nontumor tissue. We then integrated these omics datasets and asked which metabolites and lipids associate (positively or negatively) with transcript levels. Utilizing this experimental approach, we report several notable observations in tumors relative to nontumor tissue: 1) ceramides and phosphatidylserine lipids were decreased; 2) fatty acyl chain saturation was increased; 3) genes involved in complement signaling, binding and uptake of ligands by scavenger receptors, and extracellular matrix pathways were decreased; 4) and metabolites involved in amino acid and fatty acid metabolism were decreased. We also report 303 novel transcript-metabolite associations (117 gene-metabolite; 186 gene-lipid), thereby linking gene expression patterns to metabolic perturbations during HCC cancer progression in paired human tissue.

One of the key objectives of this manuscript was to determine which lipid species was most affected across tumor and adjacent nontumor tissue. Utilizing binomial analysis of lipid enrichment, we observed up to 3-fold reductions across all 22 ceramide lipid species measured regardless of acyl chain length or saturation. Ceramides are bioactive lipids important for a variety of biological processes, including cancer cell growth, proliferation, and survival (46). This finding is consistent with an independent report showing all ceramide species were two-fold lower in HCC tissue relative to nontumor tissue (47). Steady-state tissue ceramide levels can be reduced by a variety of mechanisms, including reductions in both *de novo* synthesis and catabolism of complex sphingolipids, and by accelerated ceramide turnover via ceramidases. In an attempt to explain the low levels of ceramides in tumor tissue, we first assessed the expression of genes encoding enzymes involved in *de novo* synthesis. We observed significant reductions in *SPTLC3* and 3-ketodihydrosphingosine reductase (*KDSR*) in tumor tissue, while no changes were observed in *SPTLC1, 2*, ceramide synthases (*CERS2, 4, 5, 6*), or dihydroceramide desaturases (*DEGS1, 2*). With regards to the breakdown of complex sphingolipids, such as sphingomyelin, we did observed reductions in sphingomyelin phosphodiesterases 1 (*SPMD1*) and *SMPD3* in tumors, while no changes were observed in *SPMD2, 4,* or *5*. Despite these reductions, total sphingomyelin levels were similar across tumor and adjacent nontumor tissue. Further, only N-acylsphingosine amidohydrolase 1 (*ASAH1*) was significantly reduced in tumors while neutral ceramidase (encoded by the *ASAH2* gene) and *ASAH2B* were unchanged. Taken together, these data suggest that reductions in *de novo* synthesis contribute, in part, to low ceramide levels observed in tumors relative to nontumor tissue.

While marked reductions in total ceramide levels in HCC tumors have been observed herein and by others (47), the association of serum ceramides with HCC appears to depend on chain length and degree of saturation. For instance, plasma CER(d18:1/20:1) has been shown to positively associate with tumor burden, inflammation, and shorter recurrence-free survival (RFS), while CER(d18:1/22:1) was associated with longer RFS (48). Other reports have demonstrated that all ceramide species accumulate in patients with cirrhosis and HCC relative to those without HCC (49). Thus, understanding the relationship between tissue and serum ceramide levels across HCC etiologies and disease stage is imperative.

Lipid saturation has been shown to associate with liver disease severity (50–54). For instance, patients with MASLD exhibited reductions in PUFA-triglycerides (50), and stepwise reductions in PUFA-enriched triglycerides were observed in livers from healthy controls to patients with MASH (54). Conversely, liver triglyceride saturation is increased and positively associates with increasing proton density fat fraction in adults with MASLD (51, 53). This increase in fatty acid saturation across triglyceride and phospholipid pools were also observed in HCC tumors tissue. In this study, we show selective increases in saturated and monounsaturated fatty acid-enriched triglycerides, phosphatidylcholine, and in NEFAs. This is consistent with a previous report using mass spectrometry imaging techniques to show MUFA-enriched lipids accumulate in mouse and human HCC tumors, and associate with mouse models of accelerated hepatocyte proliferation (52). Complimentary to the study by Hall et al. (52), we also observed an association of tumor MUFAs with elevations in stearoyl-CoA desaturase (SCD) transcript levels, as well as reductions in fatty acid catabolism and oxidative pathways in tumors versus nontumor tissue.

One objective of this manuscript was to detect associations of metabolite and lipid abundances with transcript levels in an attempt to identify new pathways controlling these metabolic disturbances in tumors. One of the transcripts that associated with several lipids and metabolites was that of *GMNN,* a highly oncogenic gene that is commonly amplified (frequency of ∼30%) in primary HCC (55). *GMNN* transcripts were highly associated (negatively) with lyso/phosphatidylinositol (LPI18:0, PI18:0_22:5), ceramide lipids (Cerd42:0, Cer18:0;2O/23:0, Cerd43:2), and tryptophan metabolites (3-hydroxyanthranilic acid). Notably, circulating tryptophan levels have been associated with plasma myo-inositol levels in young adults (56). Understanding the mechanisms by which *GMNN* controls DNA replication via the intersection of these metabolites may offer therapeutic benefit in HCC.

Another transcript of interest to our group was that of angiopoietin like 3 (*ANGPTL3*). Genetic loss-of-function variants in *ANGPTL3* are significantly associated with reduced cardiovascular disease (57), and antibody-mediated inhibition of *ANGPTL3* has been shown to reduce apoB-containing lipoproteins in mice and humans (57, 58). However, liver-specific antisense oligonucleotide-mediated inhibition of *ANGPTL3* led to increased hepatotoxicity (via ALT and AST levels) and increased hepatic fat content (59). In our analysis, human steatohepatitic tumors exhibited significantly lower *ANGPTL3* transcript levels (∼3.5 fold) as compared to paired nontumor tissue. In addition, *ANGPTL3* positively associated (Spearman>0.90) with MUFA- and linoleic acid-enriched phospholipids, including PC16:0_18:2, PC34:2, PE16:1_18:1, and PE34:2. Independent and supporting studies have also shown lower *ANGTPL3* expression in HCC tissues, and forced ectopic expression of *ANGPTL3* re-sensitizes sorafenib-resistant cells to sorafenib via modulation of carnitine palmitoyltransferase 1A-mediated fatty acid oxidation (60). Further, downregulation of *ANGPTL3* via siRNA in human hepatocytes increases neutral lipid accumulation via reductions in fatty acid oxidation (61). Taken together, therapeutic approaches to target circulating versus hepatic ANGPTL3 need to be carefully considered when assessing CVD versus HCC risk reduction.

It is important to acknowledge limitations in our analyses. The primary limitation is the small sample size. Given significant advancements in imaging modalities and noninvasive treatment options, HCC tissue resection is now only recommended in very early stage (Barcelona-Clinic Liver Cancer Staging System: BCLC-0) and in single intrahepatic early stage (BCLC-A) disease (62), thereby limiting the number of tissues available for research purposes. Despite the small sample size, however, a major strength of this manuscript includes the paired nature of the samples. This allows us to provide a more in-depth pairwise analyses of integrated data across omics platforms, including bulk RNA-sequencing, lipidomics, and metabolomics, with the goal of identifying novel pathways that may control cancer progression. A secondary limitation in our analysis is the amount of clinical data available on this patient cohort. Limited information regarding the stage of disease, cancer treatment(s), disease etiology, and pathological reports were available. Future studies stratifying these types of omics datasets across disease stage and/or etiology (viral vs. non-viral sources), for example, would allow for better identification of novel drivers of disease progression.

In conclusion, this manuscript was focused on identifying and describing transcript-metabolite associations in HCC, and we hope this research will lead to new pharmacological targets that may be exploited for therapeutic benefit.

## DATA AVAILABILITY

All processed data, data analyses, and analysis code have been deposited in Zenodo: https://dx.doi.org/10.5281/zenodo.18227400. All omics datasets are currently being deposited in public repositories. Additional information and requests for resources and reagents should be directed to and fulfilled by the Lead Contact, Robert N. Helsley.

## ACKNOWLEDGEMENTS

Metabolomics data were acquired and processed at the UC Davis West Coast Metabolomics Center. We would also like to thank the patients, family members, and staff for their contribution to this study.

## CONFLICTS OF INTERESTS

Conflicts of interest: nothing to report.

## SUPPLEMENTAL FIGURES AND TABLES

**Supplemental Figure 1. Experimental Design.** (**A**) An overview of the experimental design implemented in this manuscript. Image created by biorender.com.

**Supplemental Figure 2. Outlier Detection Across Omics Platforms.** (**A**-**D**) Sina plots of sample median ICI-Kt correlations across the following platforms: (**A**) RNA-sequencing, (**B**) lipidomics, (**C**) primary metabolism, and (**D**) biogenic amines and small molecule metabolites. Color indicates disease status (purple=tumor; green=nontumor) and shape indicates outlier status (triangle=outlier).

**Supplemental Figure 3. Binomial Lipid Enrichment of Phosphatidylcholine and NEFAs.** (**A**) Total number of phosphatidylcholine lipids as a function of the total number of carbons (x-axis; left) or the total number of double bonds (x-axis; right). (**B**) The total number of NEFAs as a function of the total number of carbons (x-axis; left) or the total number of double bonds (x-axis; right). Number of lipids increased (in red) or decreased (in blue) in tumors. NEFAs=nonesterified fatty acids.

**Supplemental Figure 4. Heatmap of Gene-Lipid Correlations.** (**A**) An enlarged heatmap (from **Fig. 4A**) of all 5941 gene-lipid associations. X-axis=lipid clustering (i.e. metabolite); Y-axis=gene clustering. Spearman correlation coefficient ranging from -1 (blue) to +1 (brown).

**Supplemental Figure 5. Heatmap of Gene-Metabolite Correlations.** (**A**) An enlarged heatmap (from **Fig. 6A**) of all 2800 gene-metabolite associations. X-axis=metabolite clustering (i.e. biogenic amines + primary metabolites); Y-axis=gene clustering. Spearman correlation coefficient ranging from -1 (blue) to +1 (brown).

**Supplemental Figure 6. Enrichment Analyses of Gene-Metabolite Cluster 1.** (**A**) Enrichment analyses of transcriptomic cluster 1 (via gene ontology) with metabolite cluster 1 (via reactome). X-axis=metabolite clustering (i.e. biogenic amines + primary metabolites); Y-axis=gene clustering. Spearman correlation coefficient ranging from -1 (blue) to +1 (brown).

**Supplemental Table 1.** Human Samples Analyzed Across Omics Platforms.

**Supplemental Table 2.** Differentially Expressed Genes In Tumor vs. Adjacent Nontumor Tissue

**Supplemental Table 3.** Gene Pathway Enrichment Analysis Increased in Tumors

**Supplemental Table 4.** Gene Pathway Enrichment Analysis Decreased in Tumors

**Supplemental Table 5.** Differential Lipid Abundance in Tumor versus Adjacent Nontumor Tissue

**Supplemental Table 6.** Binomial Lipid Enrichment Analysis

**Supplemental Table 7.** Gene-Lipid Correlations in Tumor vs. Adjacent Nontumor Tissue

**Supplemental Table 8.** Heatmap Cluster Enrichment Analysis of Gene-Lipid Correlations

**Supplemental Table 9.** Differential Metabolite Abundance in Tumor vs. Adjacent Nontumor Tissue

**Supplemental Table 10.** Binomial Metabolite Enrichment Analysis

**Supplemental Table 11.** Gene-Metabolite Correlations in Tumor vs. Adjacent Nontumor Tissue

**Supplemental Table 12.** Heatmap Cluster Enrichment Analysis of Gene-Metabolite Correlations

## ABBREVIATIONS

(HCC): Hepatocellular carcinoma
(MASLD): Metabolic dysfunction-associated steatotic liver disease
(SH-HCC): Steatohepatitic Hepatocellular Carcinoma
(MASH): Metabolic dysfunction-associated steatohepatitis
(MetALD): MASLD with increased alcohol intake
(H&E): Hematoxylin and eosin
(MTBE): Methyl-tert-butyl ether
(LC-HRMS/MS): Reverse-phase liquid chromatography-high resolution tandem mass spectrometry
(HILIC) LC-HRMS/MS: Hydrophilic interaction chromatography
(GC-TOFMS): Gas chromatography-time-of-flight mass spectrometry
(BMI): Body mass index
(NEFAs): Nonesterified Fatty Acids

## Notes

**FUNDING SOURCES:** This work was supported in part by the National Institute of Health (K01DK128022, R01DK139147), an American Heart Association Career Development Award (23CDA1051959), and an American Cancer Society Award (IRG2215234) to R.N.H. Research reported in this manuscript was also supported by the Biospecimen Procurement & Translational Pathology Shared Resource Facility of the University of Kentucky Markey Cancer Center (P30CA177558).

### Competing Interest Statement

The authors have declared no competing interest.

### Funding Statement

This study was funded by, in part, by the National Institute of Health (K01DK128022, R01DK139147), an American Heart Association Career Development Award (23CDA1051959), and an American Cancer Society Award (IRG2215234) to R.N.H. Research reported in this manuscript was also supported by the Biospecimen Procurement & Translational Pathology Shared Resource Facility of the University of Kentucky Markey Cancer Center (P30CA177558).

### Author Declarations

Patients undergoing surgical resection of primary HCC at the University of Kentucky (UK) were consented for tissue donation under institutional review board protocols #44026 and #70872.

