## Supplemental Figures for "An Integrated Multi-omic Analysis Reveals Novel Gene-Metabolite Relationships in Human Steatohepatitic Hepatocellular Carcinoma"

**Supplementary Figure 1**

**A.**

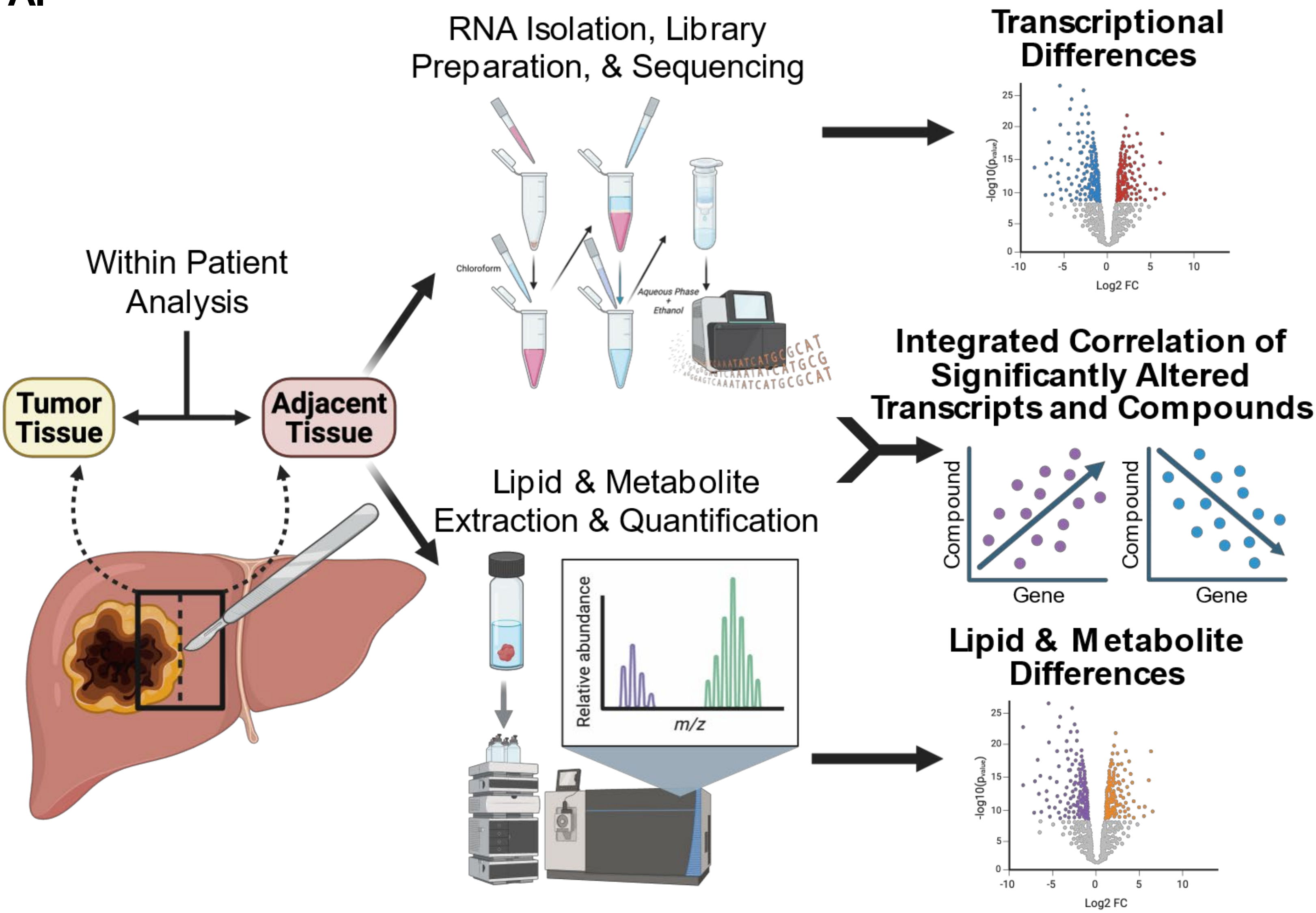

**Supplementary Figure 2**

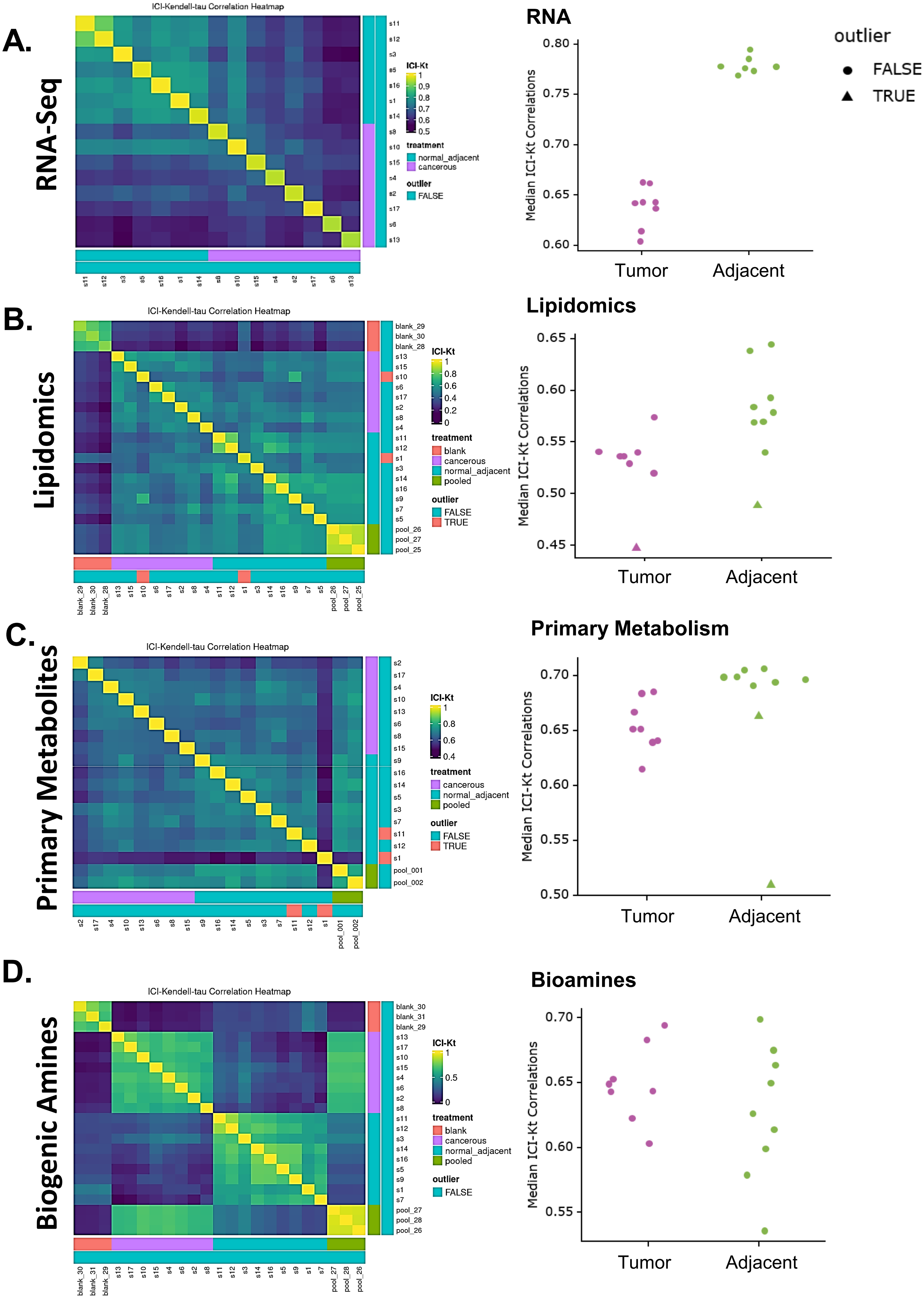

Supplementary Figure 3

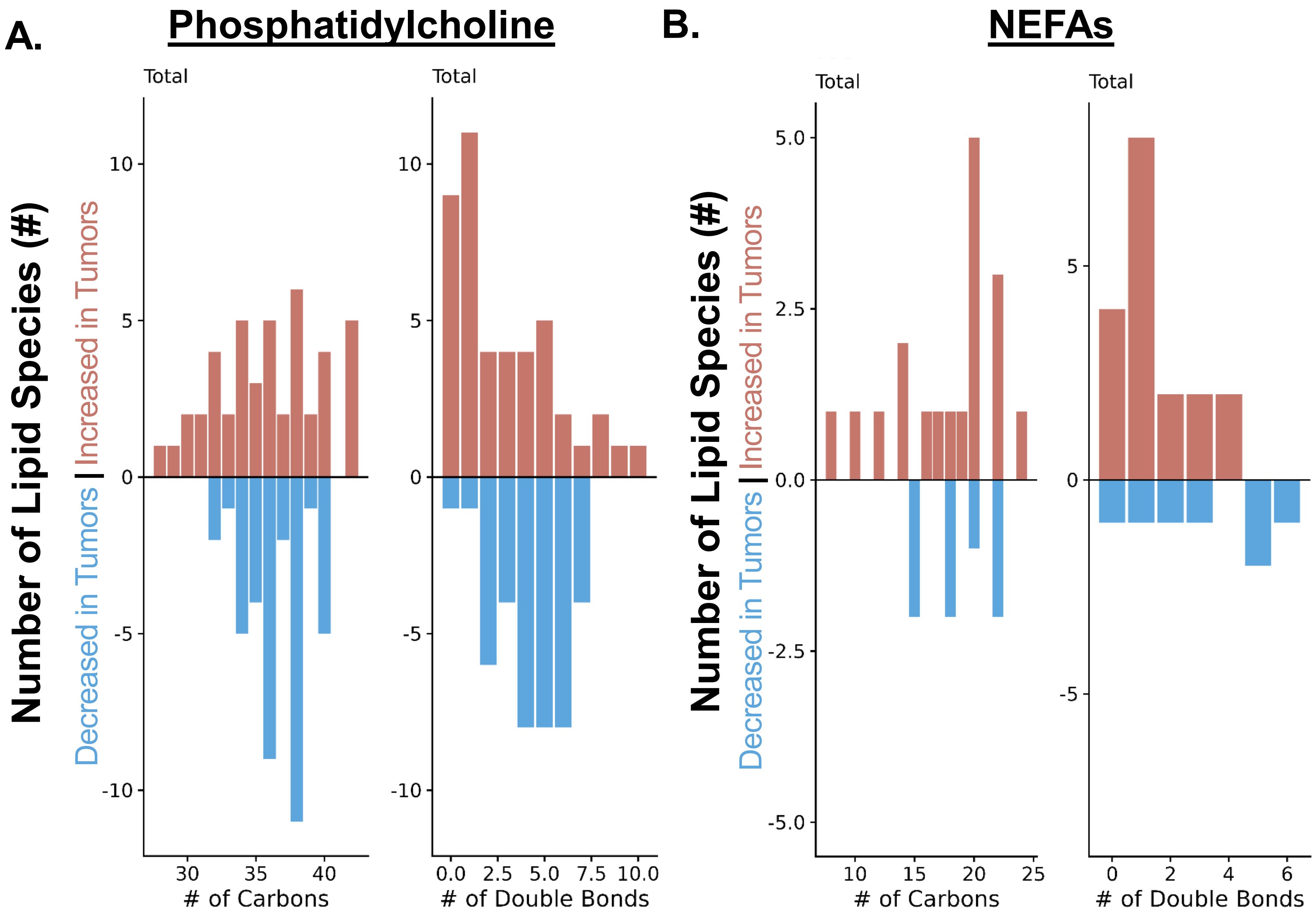

### Supplementary Figure 4

## A.

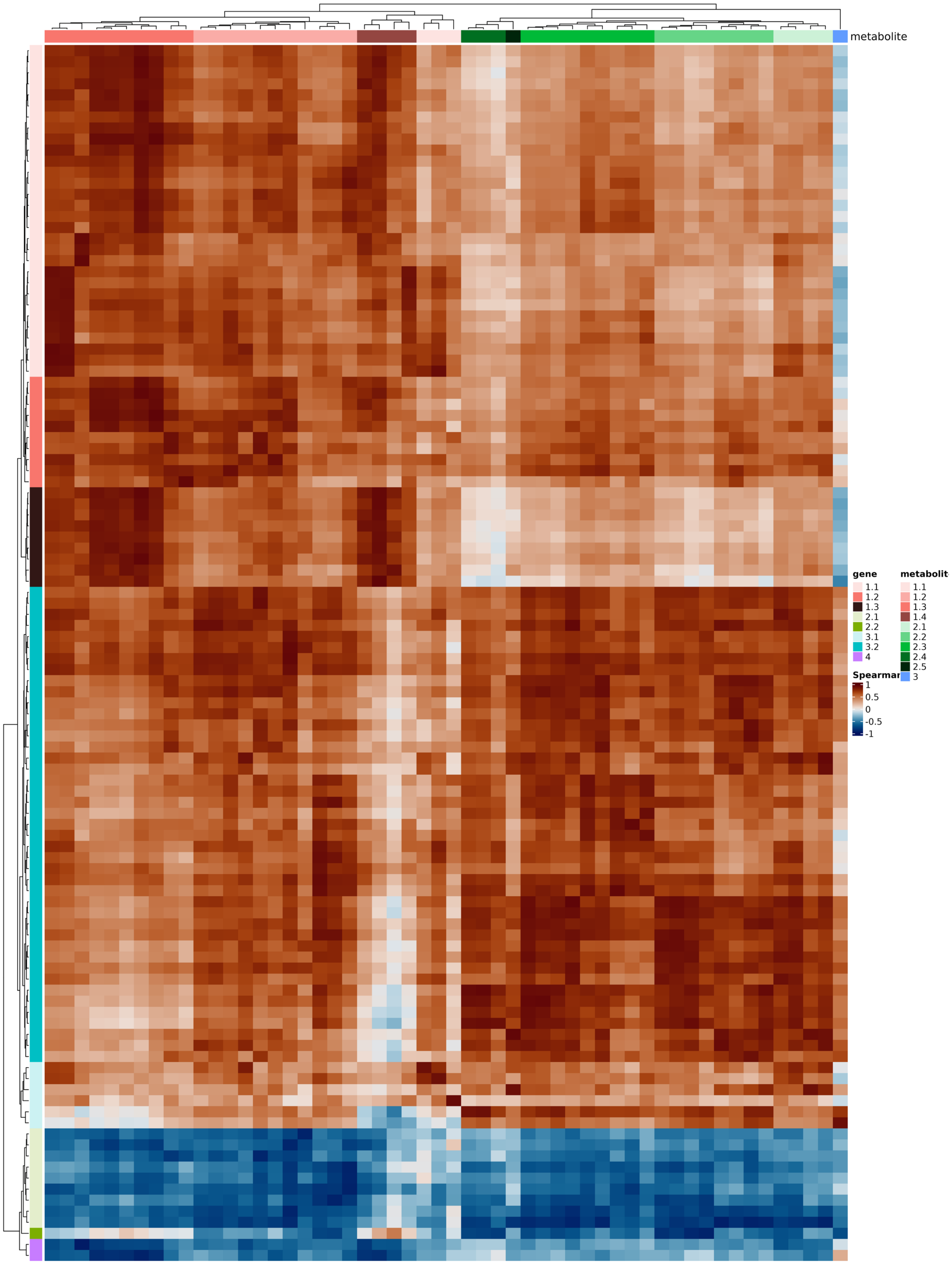

#### Supplementary Figure 5

**A.**

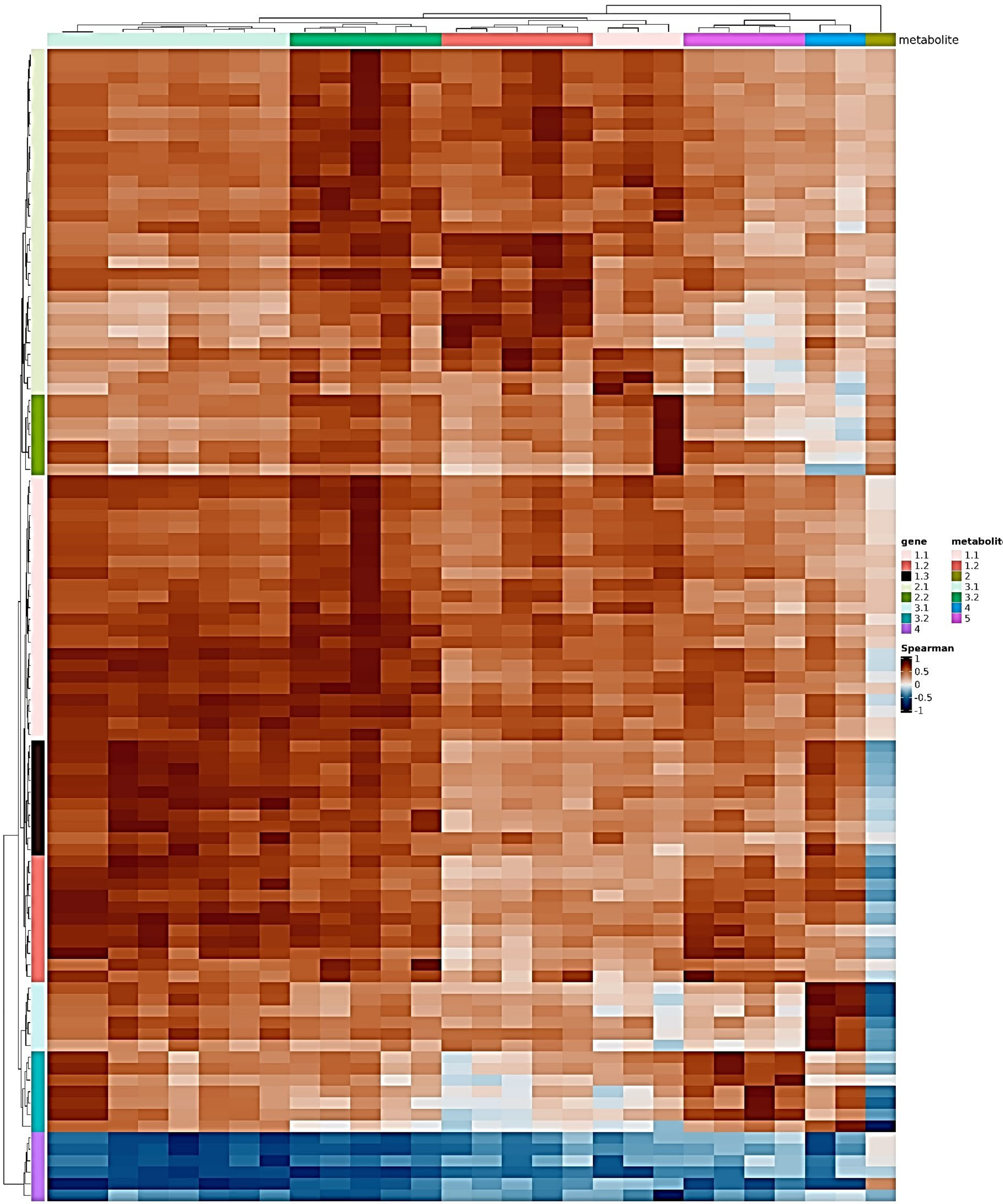

### Supplementary Figure 6

**A.**

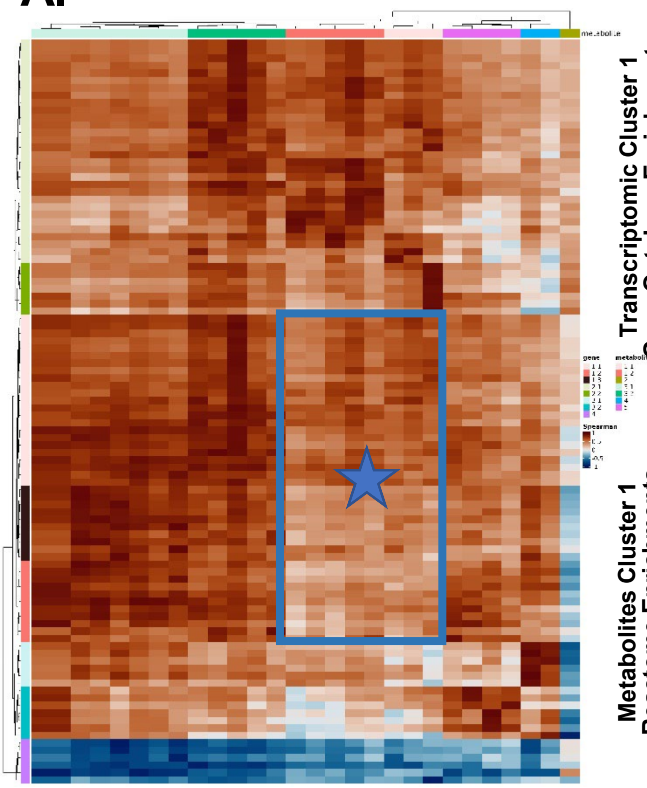

**Transcriptomic Cluster 1 Gene Ontology Enrichments**

| description | p | padjust | expected | counts | significant | odds | ID |
|---|---|---|---|---|---|---|---|
| BP:organic acid catabolic process | 9.25 × 10−14 | 7.56 × 10−10 | 0.30 | 6 | 33 | 25.85 | GO:0005201 |
| BP:carboxylic acid catabolic process | 5.77 × 10−14 | 7.56 × 10−10 | 0.85 | 8 | 33 | 12.44 | GO:0062023 |
| BP:monocarboxylic acid catabolic process | 5.39 × 10−13 | 2.94 × 10−9 | 1.03 | 8 | 33 | 10.17 | GO:0030312 |
| BP:small molecule catabolic process | 7.92 × 10−12 | 3.24 × 10−8 | 1.03 | 8 | 33 | 10.17 | GO:0031012 |

**Metabolites Cluster 1 Reactome Enrichments**

| description | p | padjust | expected | counts | significant | odds | ID |
|---|---|---|---|---|---|---|---|
| POLB-Dependent Long Patch Base Excision Repair | 2.23 × 10−3 | 2.97 × 10−2 | 0.1 | 2 | 8 | Inf | R-HSA-110362 |
| Resolution of AP sites via the multiple-nucleotide patch replacement pathway | 2.23 × 10−3 | 2.97 × 10−2 | 0.1 | 2 | 8 | Inf | R-HSA-110373 |
| PIP3 activates AKT signaling | 2.23 × 10−3 | 2.97 × 10−2 | 0.1 | 2 | 8 | Inf | R-HSA-1257604 |
| Signaling by TGF-beta Receptor Complex | 2.23 × 10−3 | 2.97 × 10−2 | 0.1 | 2 | 8 | Inf | R-HSA-170834 |
