## Supplemental Table 1 for "An Integrated Multi-omic Analysis Reveals Novel Gene-Metabolite Relationships in Human Steatohepatitic Hepatocellular Carcinoma"

**Supplemental Table 1.** Human Samples Analyzed Across Omics Platforms.

| **Patient** | **Sample #** | **Tissue** | **RNA-seq** | **Lipidomics** | **1° Metabolism^#^** | **Bioamines^#^** | **Correlation** |
| --- | --- | --- | --- | --- | --- | --- | --- |
| 1 | s1 | Adjacent Nontumor | Yes | Outlier | Outlier | Yes | N/A |
| 1 | S2 | Tumor | Yes | Yes | Yes | Yes | Yes |
| 2 | s3 | Adjacent Nontumor | Yes | Yes | Yes | Yes | Yes |
| 2 | s4 | Tumor | Yes | Yes | Yes | Yes | Yes |
| 3 | s5 | Adjacent Nontumor | Yes | Yes | Yes | Yes | Yes |
| 3 | s6 | Tumor | Yes | Yes | Yes | Yes | Yes |
| 4 | s7 | Adjacent Nontumor | No* | Yes | Yes | Yes | N/A |
| 4 | s8 | Tumor | Yes | Yes | Yes | Yes | Yes |
| 5 | s9 | Adjacent Nontumor | No* | Yes | Yes | Yes | N/A |
| 5 | s10 | Tumor | Yes | Outlier | Yes | Yes | N/A |
| 6 | s11 | Adjacent Nontumor | Yes | Yes | Outlier | Yes | N/A |
| 6 | s12 | Adjacent Nontumor | Yes | Yes | Yes | Yes | Yes |
| 6 | s13 | Tumor | Yes | Yes | Yes | Yes | Yes |
| 7 | s14 | Adjacent Nontumor | Yes | Yes | Yes | Yes | Yes |
| 7 | s15 | Tumor | Yes | Yes | Yes | Yes | Yes |
| 8 | s16 | Adjacent Nontumor | Yes | Yes | Yes | Yes | Yes |
| 8 | s17 | Tumor | Yes | Yes | Yes | Yes | Yes |
| ***Correlations were only completed on samples with complete datasets*** | | | | | | | |
| ***^#^*UC West Coast Metabolomics Platforms** | | | | | | | |
| Paired analyses were performed | | | | | | | |
| Paired analyses were not performed | | | | | | | |
| *No = RNA quality was too low to perform RNA-sequencing | | | | | | | |
